# The phase angle is associated with upper arm muscle circumference but not with sarcopenia in continuous ambulatory peritoneal dialysis patients

**DOI:** 10.1101/2022.07.10.22277470

**Authors:** Ye Chen, Jinlan Wu, Lei Ran, Dan Yu, Xi Chen, Maodong Liu

## Abstract

**Aims:** Limited studies on phase angle (PhA) and sarcopenia in continuous ambulatory peritoneal dialysis (CAPD) patients. We want to explore the association between PhA and sarcopenia and clarify its significance and value in diagnosing sarcopenia.

**Methods:** We included 87 CAPD patients. We measured the PhA and body composition by bioelectrical impedance analysis. All patients had their handgrip strength(HGS) measured. Then, we divided patients into the sarcopenia (n=25) group and non-sarcopenia(n=62) group according to the sarcopenia diagnostic strategy formulated by the Asian Working Group for Sarcopenia.

**Results:** Univariate analysis shows PhA is positively associated with HGS, skeletal muscle mass (SMM), and Upper arm muscle circumference (AMC). After adjustments for sex, age, diabetes, BMI, extracellular water ratio(E/T), extra water (OH), serum creatinine, total kt/v, and residual kt/v, PhA remains correlated to HGS and AMC, but not to SMM. In the multivariate logistic model, PhA is not correlated to sarcopenia. The AUROC of PhA for sarcopenia is 0.76(95% CI, 0.65-0.86, P <0.01, fig 3). The optimal cut-off value is identified as ≤5.25(sensitivity 74%, specificity 76%).

**Conclusion:** This study illustrates that older age and higher total kt/v are risk factors for sarcopenia. PhA is positively associated with HGS and AMC but not with sarcopenia in CAPD patients. These results suggest that PhA can predict muscle mass and strength in CAPD patients, but its diagnosis value for sarcopenia needs more studies.

## Introduction

Sarcopenia is the loss of muscle mass, strength, or muscle function, which was initially thought to be an age-related disease[1]. In contrast, recent studies showed that some inflammatory diseases, such as tumors and organ failure, could increase the risk of sarcopenia[2]. In addition, sarcopenia is one of the most important complications in patients undergoing peritoneal dialysis due to micro-inflammatory state, inadequate caloric intake, uremic toxin accumulation, volume overload, and peritoneal dialysis process [3], with a prevalence of 4%-15.5%[4, 5]. Several studies demonstrated that sarcopenia was associated with physical disabilities, higher mortality, cardiovascular events, and falls in chronic kidney disease (CKD). In 2015, a prospective analysis of hemodialysis (HD) patients presented a higher risk of disabilities in sarcopenia than non-sarcopenia group[6]. Yuka et al.[7]enrolled 119 PD patients and found that sarcopenia was associated with a higher risk of mortality (survival mortality rate per 500 days: 0.667 in the sarcopenia group vs. 0.971 in the non-sarcopenia group, p<0.001). Then, a study of elderly HD patients showed sarcopenia patients presented a higher risk of falls than non-sarcopenia ones. According to a prospective observation study[8], sarcopenia was a strong predictor of death (HR, 6.99; 95% CI, 1.84 to 26.58; p = 0.004) and cardiovascular events (HR, 4.33; 95% CI, 1.51 to 12.43; p = 0.006). In addition to this, a multicenter analysis confirmed a correlation between sarcopenia and a high risk of hospitalization in HD patients(RR,2.07, 95%CI, 1.48-2.88, P<0.001). In summary, the use of easily accessible and cheaper instruments to test the nutritional status of patients is extremely important for the diagnosis and early prevention of sarcopenia in clinical practice.

Bioelectrical impedance analysis (BIA) is quick and relatively noninvasive to analyze the body composition of the human body, which can measure muscle, fat, and water separately[9]. Thus, it has been widely used for body composition measurement. BIA is more valuable in chronic kidney disease due to the volume overload in those patients. Recent guidelines recommend BIA could be used to estimate muscle mass in diagnosing sarcopenia[2]. PhA was an indicator measured by BIA and has been gaining attention because it reflected the cellular function and the distribution of intracellular and extracellular water[10]. Several studies found it correlated to muscle strength and function[11, 12]. Thus, the current evidence encouraged research on the use of PhA in the nutrition care process and diagnosis, particularly in sarcopenia. Many studies have explored the relationship between sarcopenia and PhA in different diseases, including patients undergoing PD. However, there are few studies on Chinese CAPD patients. After all, the phase angle value varies from one race to another[13].

In clinical practice, there are many CAPD patients in our nephrology clinic. Exploring easy ways to find sarcopenia in these patients draws our attention. This study wants to explore the association between PhA and sarcopenia in Chinese CAPD patients. Meanwhile, we want to find an optimal cut-off value of sarcopenia diagnostic in these patients.

## Materials and methods

### Study design and patient recruitment

This cross-sectional study was carried out between August 2021 and March 2022. It evaluated CAPD patients in the Peritoneal Dialysis Clinic of The Third Hospital of Hebei Medical University, Shijiazhuang, Hebei Province. Inclusion criteria were patients aged >18 years, on stable dialysis>3 months, and with regular follow-up. Patients with acute infection at any site (respiratory, digestive, urinary), acute cardiovascular disease, malignancy, active liver disease, tuberculosis, AIDS, syphilis, and other systemic diseases, surgery, trauma, or disability resulting in deficient grip strength and body composition analysis data were excluded. The Ethics Committee approved the study protocol of The Third Hospital of Hebei Medical University **(number:20210931)**. All subjects signed an informed consent form.

### Anthropometric measurements

The Clinical nutrition test analyzer (AINST 2018, Taizhou city, Jiangsu province, China) records the patient’s body weight with a precision of 1g (we will subtract the weight of peritoneal fluid when measuring). Height was measured by a Portable height measuring instrument with a precision of 0.1cm.

### Body composition analysis and phase angle

Determination of body composition, PhA and AMC use a clinical nutrition test analyzer based on the BIA method. Subjects were asked to fast for at least 8 hours and empty their urine and stool in the early morning of the test. Measurement method: Clean the electrode contact surface of the instrument with an alcohol cotton ball, measure the height and weight of the subject; the subject is barefoot, wipe the soles of both feet and palms of both hands with an alcohol cotton ball, make both feet and hands contact the silver electrode contact surface of the instrument to the maximum extent, and introduce the current. Start to measure and read total water, extra water (OH), protein, skeletal muscle mass (SMM), body fat, appendicular skeletal muscle mass index (ASMI), extracellular water ratio(E/T), AMC, PhA, and other data.

### Baseline variables

Sociodemographic backgrounds were obtained via face-to-face interviews, including name, sex, and age. Clinical data, including dialysis vintage, blood pressure, and year diagnosed with ESRD, come from electronic or paper cases from our center. Routine laboratory tests, including high-sensitive C-reactive protein(Hs-CRP), serum hemoglobin, and so on, were collected from medical records. Blood samples are collected on an empty stomach on the morning of the day of the body composition measurement.

### Grip strength

Handgrip strength (HGS) is measured by a Grip Strength meter (CAMRY, MODEL: EH101). We ask subjects to turn their palms inward with the dial facing outward. Their bodies stand upright, and their arms hang down naturally. The grip meter should not contact their bodies and clothes. Then, use the force of the muscle group of the main hand to measure three times and take the average value.

### Total kt/V_urea_ Assessment

A 24 h urine sample and dialysis fluid specimen were collected, and fasting venous blood was taken in the morning of the following day. Kt/V_urea_ is calculated. At the same time, professionals checked and recorded Dialysis protocol, urine volume, and ultrafiltration volume for the day.

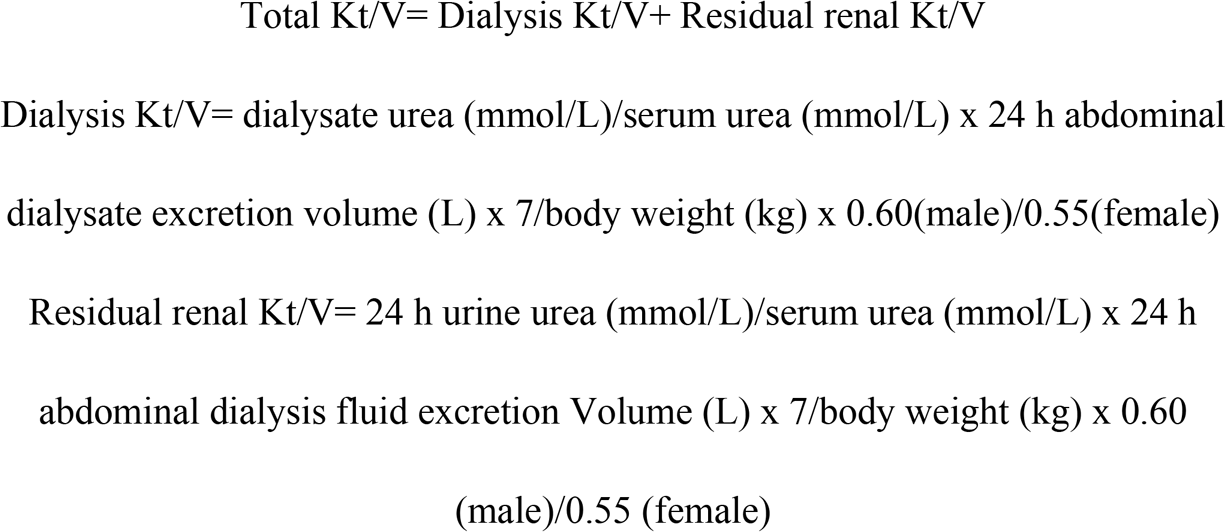

### Sarcopenia Diagnosis

According to the Asian sarcopenia diagnostic strategy formulated by the Asian Working Group for Sarcopenia (AWGS), the subjects should meet the following two criteria of the BIA test:1. appendicular skeletal muscle mass index (ASMI)=appendicular skeletal muscle(kg)/height (m^2^): Males<7.0kg/m^2^, Females<5.7kg/m^2^; 2. Handgrip strength: Males<26kg, females<18kg.

### Statistical Analysis

We analyzed the data using the statistical software IBM SPSS Statistics version 26.0(SPSS Inc., Chicago, IL, United States). Continuous variables with normal distribution were presented as mean ± standard deviation (SD), whereas skewed data were presented as median (q1–q3). The number of cases (percentage) was used for categorical counts.

The Pearson product-moment correlation assesses the correlation between PhA and diagnostic indicators of sarcopenia. Logistic regression analysis is performed to assess the independent predictors of sarcopenia. A multivariate analysis is adjusted for age, total Kt/V, residual kidney Kt/V, basal metabolic, OH, E/T and serum creatinine.

We use a receiver-operating characteristics (ROC) curve analysis to determine diagnostic accuracy and the cut-off point for PhA to detect sarcopenia. The area under the curve (AUC) indicates the discriminative power of the test. The Youden index calculated the best cut-off value in the AUC. P < 0.05 was considered a statistically significant difference.

## Results

### 1. The basic information of peritoneal dialysis patients

A total of 102 patients on peritoneal dialysis came to our outpatient clinical from September 2021 to March 2022. Seven patients had been on peritoneal dialysis for less than three months. Four patients refused to be enrolled, two patients had hemiplegia of the limbs due to old cerebral infarction, and two were younger than 18 years old. Eighty-seven patients were finally enrolled, including 43 males(49.43%) and 44 females(50.57%), with a mean age of 51.3±12.43 years, and 25 patients were diagnosed with sarcopenia. (Table 1)

**Table 1.** Comparison of general condition, grip strength, body composition analysis and hematological indices in sarcopenic and non-sarcopenic abdominal dialysis patients

| Variables | Total | Sarcopenia group | Non-sarcopenia group | t/z/$\chi^2$ value | P-value |
| --- | --- | --- | --- | --- | --- |
| **Baseline variables** |  |  |  |  |  |
| Age | $51.3 \pm 12.43$ | $62.12 \pm 9.20$ | $46.94 \pm 10.83$ | 6.169 | <0.01 |
| Sex | Male 43 (49.43%) | Male 10 (40%) | Male 33(53.23%) | 1.247<sup>a</sup> | 0.264 |
| DM | Yes 17 (19.54%) | Yes 7 | Yes 10 | 1.597<sup>a</sup> | 0.2 |
| SBP | $141.03 \pm 11.59$ | 141(130-150) | $141.5(132.75-150)$ | -0.028 | 0.98 |
| DBP | 90(87-99) | 90(86-96.5) | 90(87.75-99) | -0.557 | 0.58 |
| Dialysis age<br>(month) | 34(17-71) | 21(16-72) | 37.5(17.75-70.25) | -0.685 | 0.493 |
| **Bioelectrical impedance analysis** |  |  |  |  |  |
| BFP (%) | $26.64 \pm 9.54$ | $29.31 \pm 11.14$ | $25.56 \pm 8.68$ | 1.676 | 0.097 |
| WHR | $0.82 \pm 0.10$ | $0.83 \pm 0.11$ | $0.82 \pm 0.09$ | 0.339 | 0.74 |
| AC (cm) | $28.93 \pm 3.37$ | $28.11 \pm 3.21$ | $29.26 \pm 3.40$ | -1.449 | 0.151 |
| AMC (cm) | $23.44 \pm 2.48$ | $22.69 \pm 2.16$ | $23.75 \pm 2.55$ | -1.830 | 0.071 |
| BMI (kg/m<sup>2</sup>) | 23.9(20.8-26.6) | 24.4(20.95-25.85) | 23.6(20.73-26.73) | -1.88 | 0.85 |
| BMR (kcal) | 1368.61(1206.85-1598.16) | 1279.16(1198.48-1394.91) | 1445.35(1203.25-1617.87) | -1.988 | 0.047 |
| PhA (°) | 5.4(4.5-5.8) | 4.5(4.05-5.3) | 5.45(5.2-5.9) | -3.786 | <0.018 |
| Cell mass(kg) | 28.91(24.36-35.62) | 27.23(23.83-29.99) | 31.65(24.46-37.08) | -2.22 | 0.027 |
| LBM (kg) | 46.23(38.74-56.86) | 42.09(38.355-47.45) | 49.79(38.58-57.77) | -1.988 | 0.047 |
| VFA (cm²) | 72.89(60.13-106.73) | 96.59(62.71-138.84) | 72.09(53.87-101.53) | -1.505 | 0.132 |
| BMC (kg) | 2.73(2.34-3.19) | 2.66(2.36-2.84) | 2.92(2.32-3.33) | -1.674 | 0.094 |
| Protein (kg) | 8.73(7.35-10.75) | 8.22(7.19-9.06) | 9.55(7.38-11.19) | -2.214 | 0.027 |
| SMM (kg) | 24.32(20.18-30.43) | 22.79(19.70-25.31) | 26.82(20.27-31.77) | -2.223 | 0.026 |
| Muscle mass(kg) | 43.45(36.11-53.48) | 39.64(35.86-44.61) | 46.93(36.20-54.54) | -2.026 | 0.043 |
| ECW (L) | 13.59(11.33-16.81) | 12.43(11.45-14.47) | 14.63(11.11-17.11) | -1.459 | 0.145 |
| ICW (L) | 20.18(17.01-24.87) | 19.01(16.64-20.94) | 22.10(17.08-25.89) | -2.214 | 0.027 |
| Total water(L) | 34.05(28.34-42.07) | 30.92(28.22-34.99) | 36.86(28.28-42.73) | -1.928 | 0.054 |
| OH(L) | 1.21(0.75-1.9) | 1.4(1.21-1.95) | 1.10(0.70-1.88) | -2.303 | 0.021 |
| E/T | 0.4(0.39-0.41) | 0.4(0.4-0.42) | 0.39(0.39-0.4) | -3.7 | <0.01 |
| **Routine laboratory tests** |  |  |  |  |  |
| Dialysis dose(ML) | 8552(6280-9090) | 8420(6145-8930) | 8617.5(6318.25-9100) | -1.013 | 0.311 |
| Total Kt/V | 1.80±0.29 | 1.93±0.35 | 1.74±0.25 | 2.791 | <0.01 |
| Residual kidney kt/V | 0(0-0.473) | 0.236(0-0.80) | 0(0-0.30) | -2.528 | 0.011 |
| Dialysis kt/V | 1.54(1.34-1.80) | 1.601(1.27-1.84) | 1.53(1.35-1.80) | -0.01 | 0.996 |
| Hb | 110(103-120) | 110(103-118.5) | 109.5(102.75-122) | -0.028 | 0.978 |
| Hs-CRP | 3.52(1.78-9.87) | 3.2(1.58-6.97) | 3.6(1.93-11.27) | -1.018 | 0.309 |
| ALB | 38.61(35.82-41.47) | 36.66(33.83-39.08) | 39.09(36.05-42.63) | -2.762 | 0.006* |
| TIBC | 44.05(36.96-50.5) | 43.2(35.68-47.84) | 44.63(37.34-52.82) | -1.116 | 0.264 |
| FER | 194.44(77.03-407.69) | 153.62(80.20-367.28) | 197.99(75.47-425.19) | -0.347 | 0.729 |
| SI | 11.26(9.29-14.5) | 11.61(8.66, 13.81) | 11.12(9.52-15.34) | -0.657 | 0.511 |
| TS | 0.26(0.21-0.34) | 0.28(0.23, 0.33) | 0.25(0.20-0.34) | -0.919 | 0.358 |
| Cr | 984.06(812.54-1187.94) | 806.37(621.86-959.23) | 1074.07(897.06-1287.97) | -4.596 | <0.01 |
| BUN | 19.88(16.55-23.05) | 19.91±4.92 | 20.76±4.81 | -0.736 | 0.464 |
| HCO3- | 25.00±2.71 | 24.43±2.49 | 25.23±2.78 | -1.252 | 0.214 |
| PA | 369.7(323.2-414.4) | 326.9(289.65-383.7) | 379.25(339.25-424.83) | -2.598 | <0.01 |
| TG | 1.45(1.13-2.32) | 1.32(1.01-2.09) | 1.55(1.15-2.33) | -1.177 | 0.239 |
| TC | 5.15±1.13 | 5.22±1.07 | 5.11±1.16 | 0.387 | 0.7 |
| HDL | 1.26(1.01-1.54) | 1.35(1.04-1.76) | 1.22(1.01-1.47) | -1.229 | 0.219 |
| LDL | 2.85±0.74 | 2.85±0.65 | 2.86±0.78 | -0.031 | 0.975 |
| VLDL | 0.68(0.51-1.19) | 0.63(0.5-2) | 0.7(0.51-1.05) | -0.403 | 0.687 |
| ALP | 75(59-107) | 76(61-102.5) | 74(58-108.5) | -0.277 | 0.782 |
| PTH | 212.2(468.3-727.4) | 338.5(192.75-541.2) | 538.55(212.5-770.28) | -1.984 | 0.047 |

DM: Diabetic Mellitus; SBP: Systolic blood pressure; DBP: Diastolic blood pressure; BFP: Body fat percentage; WHR: Waist-Hip ratio; AC: Upper arm circumference
AMC: Upper arm muscle circumference; BMR: Basal metabolic rate; PhA: Phase angle; LBM: Lean body mass; VFA: Visceral fat area; IS: Inorganic salts; BMC: Bone mineral content; SMM: Skeletal muscle mass; ECW: Extracellular water; ICW: Intracellular water; OH: Extra water; E/T: Extracellular water ratio; Hb: Hemoglobin; Glu: Blood glucose; Hs-CRP: Hypersensitive C-reactive protein; Ca2+: Serum calcium; P+: Serum phosphorus; K+: Serum potassium; Na+: Serum sodium; Cl-: Serum chloride; ALB: Serum albumin; TIBC: Total iron-binding force; FER: Ferroprotein; SI: Serum iron; TS: Transferrin saturation; Cr: Serum creatinine; UA: serum uric acid; BUN: Serum urea; HCO3-: Bicarbonate; PA: Prealbumin; TG: Triglycerides; TC: Total cholesterol; HDL: High-density lipoprotein; LDL: Low-density lipoprotein; VLDL: Very low-density lipoprotein; ALP: Alkaline phosphatase; PTH: Parathyroid hormone.

### 2. The comparison between the sarcopenia group and non-sarcopenia group

Age is significantly higher in the sarcopenia group than in the non-sarcopenia group. The total Kt/V and residual kidney Kt/V are higher in the sarcopenia group than in the non-sarcopenia group. But the total Kt/v in non-sarcopenia patients is approximately 1.7, which is the best total Kt/v for peritoneal dialysis patients. The body composition analysis shows that basal metabolic rate, PhA, cell mass, de-fatted body weight, protein, skeletal muscle, muscle mass, intracellular water, serum calcium, serum sodium, serum albumin, serum pre-albumin, serum creatinine, and parathyroid hormone (PTH) are all lower in the sarcopenia group than the non-sarcopenia group;. In contrast, excess water and E/T are higher in the sarcopenia group than in the non-sarcopenia group. (Table 1)

### 3. The relationship between PhA and diagnostic indicators of sarcopenia

Pearson correlation analysis shows a positive correlation between PhA with HGS, skeletal muscle mass (SMM), and ACM, but not with ASMI and muscle mass (Model 1). After adjustments for age, diabetes, BMI, E/T, OH, serum creatinine level, total Kt/V, and residual Kt/V, the positive correlation between PhA with HGS and muscle circumference of upper arm remained (Model 7), except for SMM, ASMI, and muscle mass (Table 2).

**Table 2.** The person correlation between PhA with Indicators related to sarcopenia

| Model<br>variables | Model 1 |  | Model 2 |  | Model 3 |  | Model 4 |  | Model 5 |  | Model 6 |  | Model 7 |  |
| --- | --- | --- | --- | --- | --- | --- | --- | --- | --- | --- | --- | --- | --- | --- |
|  | r | p | r | p | r | p | r | p | r | p | r | p | r | p |
| PhA×HGS | 0.49 | <0.01 | 0.22 | 0.04 | 0.21 | 0.06 | 0.19 | 0.08 | 0.17 | 0.14 | 0.26 | 0.02 | 0.23 | 0.048 |
| PhA×ASMI | 0.18 | 0.09 | -0.07 | 0.52 | -0.23 | 0.04 | -0.22 | 0.05 | -0.14 | 0.20 | -0.14 | 0.32 | -0.11 | 0.34 |
| PhA×muscle mass | 0.19 | 0.08 | -0.16 | 0.15 | 0.27 | 0.01 | 0.20 | 0.08 | 0.2 | 0.08 | 0.19 | 0.1 | 0.10 | 0.37 |
| PhA×SMM | 0.26 | 0.02 | -0.05 | 0.65 | 0.27 | 0.01 | 0.20 | 0.08 | 0.2 | 0.08 | 0.19 | 0.1 | 0.10 | 0.37 |
| PhA×AMC | 0.42 | <0.01 | 0.51 | <0.01 | 0.30 | <0.01 | 0.28 | 0.01 | 0.31 | <0.01 | 0.31 | <0.01 | 0.24 | 0.03 |

Model 1 Univariate correlation; Model 2 adjusted for sex, age, diabetes, and BMI; Model 3 adjusted for sex, age, diabetes, BMI, and Extracellular water ratio; Model 4: adjusted for sex, age, diabetes, BMI, Extracellular water ratio and extra water; Model5: adjusted for sex, age, diabetes, BMI, Extracellular water ratio, extra water, and serum creatinine; Model 6: adjusted for sex, age, diabetes, BMI, Extracellular water ratio, extra water, serum creatinine, and total kt/v; Model 7: adjusted for sex, age, diabetes, BMI, Extracellular water ratio, extra water, serum creatinine, total kt/v, and residual kt/v.

### 4. Univariate and Multivariate logistic regression between PhA and sarcopenia

A univariate logistic regression between PhA and sarcopenia(Table 3) shows that with PhA Q1 as the baseline level, the ORs for the risk of disease for Q2, Q3, and Q4 is 0.583(95%CI 3.15-248.78), 0.163(95%CI 1.75-152.82) and 0.036(95%CI 0.492-42.33). The Q1, Q2, Q3, and Q4 are the quartile levels of the PhA. The risk of sarcopenia decreases with increasing PhA. However, after adjustments for age, total Kt/V, residual kidney Kt/V, basal metabolic, OH, E/T, and serum creatinine, PhA is not a risk factor for sarcopenia(P>0.05). As the diagnostic criteria included HGS and ASMI, HGS and muscle-related indicators such as SMM and muscle mass were excluded from the logistic regression.

**Table 3.** Logistic Regression analysis for sarcopenia according to different variables.

| Model<br>Variables | univariate<br>Odds ration(95%CI) | p | multivariate<br>Odds ration(95%CI) | p |
| --- | --- | --- | --- | --- |
| **Phase angle** |  | 0.003 |  | 0.710 |
| Q2(ref Q1) | 0.583(3.151-248.774) | 0.003 | 3.076(0.025-372.251) | 0.646 |
| Q3(ref Q1) | 0.163(1.746-152.822) | 0.014 | 0.348(0.004-0.65) | 0.650 |
| Q4(ref Q1) | 0.036(0.492-42.33) | 0.181 | 0.123(0-68.623) | 0.516 |
| **Age** |  | <0.01 |  | 0.069 |
| Age≥45 (ref age<45) | 4.762(0.494-45.945) | 0.177 | 30.437(0.534-1707.367) | 0.096 |
| Age≥55 | 14.286(1.616-126.297) | 0.017 | 47.507(0.613-3683.278) | 0.082 |
| Age≥65 | 150(12.347-1822.25) | <0.01 | 675.960(4.919-92894.294) | 0.009 |
| **E/T(ref E/T≤0.4)** | 3.479(1.287-9.408) | 0.014 | 1.127(0.015-86.132) | 0.957 |
| **OH** |  | 0.014 |  | 0.459 |
| Q2(ref Q1) | 1.895(0.308-11.644) | 0.490 | 0.253(0.006-10.882) | 0.474 |
| Q3 | 10.800(2.003-58.224) | <0.01 | 3.294(0.042-257.569) | 0.592 |
| Q4 | 4.2(0.756-23.232) | 0.101 | 1.266(0.003-525.443) | 0.939 |
| **Basal metabolic ration** |  | 0.1 |  | 0.342 |
| Q2(ref Q1) | 2.083(0.588-7.383) | 0.256 | 19.486(0.701-541.805) | 0.080 |
| Q3 | 1.167(0.317-4.299) | 0.817 | 4.059(0.181-91.23) | 0.378 |
| Q4 | 0.25(0.044-1.417) | 0.117 | 3.547(0.038-331.756) | 0.585 |
| **Total Kt/V** |  | 0.023 |  | 0.115 |
| Total kt/v≥1.544 (ref total kt/v<1.544) | 1.333(0.26-6.828) | 0.73 | 4.705(0.081-272.491) | 0.455 |
| Total kt/v≥1.797 (ref total kt/v<1.544) | 2.25(0.482-10.504) | 0.302 | 10.714(0.169-677.935) | 0.262 |
| Total kt/v≥2.013 (ref total kt/v<1.544) | 7.2(1.635-31.712) | <0.01 | 121.131(1.660-8839.286) | 0.028 |
| **Residual kidney Kt/V(ref 无)** | 3.806(1.386-10.447) | <0.01 | 1.77(0.224-13.971) | 0.588 |
| **Serum creatinine** |  | <0.01 |  | 0.530 |
| Q2(ref Q1) | 0.287(0.082-1.01) | 0.052 | 0.927(0.057-15.11) | 0.958 |
| Q3 | 0.137(0.034-0.552) | <0.01 | 2.809(0.121-65.12) | 0.520 |
| Q4 | 0.029(0.003-0.262) | <0.01 | 0.119(0.002-6.084) | 0.289 |

The Q1, Q2, Q3, and Q4 are the quartile levels of each variable

### 5. AUROC of PhA for sarcopenia

The AUROC of PhA for sarcopenia is 0.76(95% CI, 0.65-0.86, P<0.01, fig 3).

The optimal cut-off value is ≤5.25(sensitivity 74%, specificity 76%,fig3). On the other hand, the AUROC of males is 0.85(95%CI, 0.73-0.97), and the cut-off point of PhA for males is 5.25(sensitivity 81.8%, specificity 80%,fig 1). Whereas the AUROC of females is 0.67, and the best predictive value of PhA for sarcopenia is 5.1 (sensitivity 69%, specificity 73%,fig 2), which is lower than for men.

**Fig 1.**
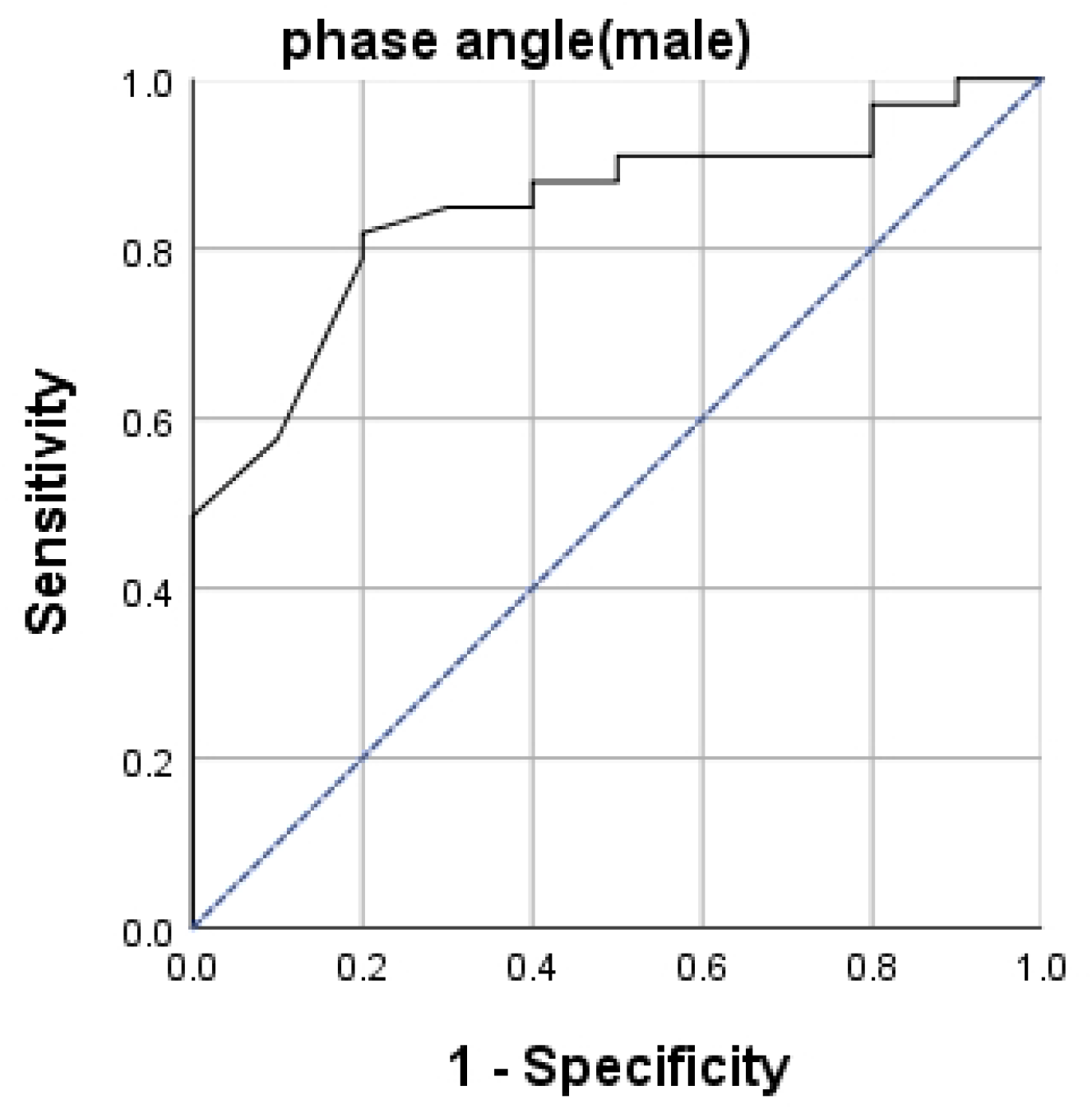
The ROC curve of PhA for sarcopenia in males, the AUCROC is 0.85(p<0.01).

**Fig 1.**
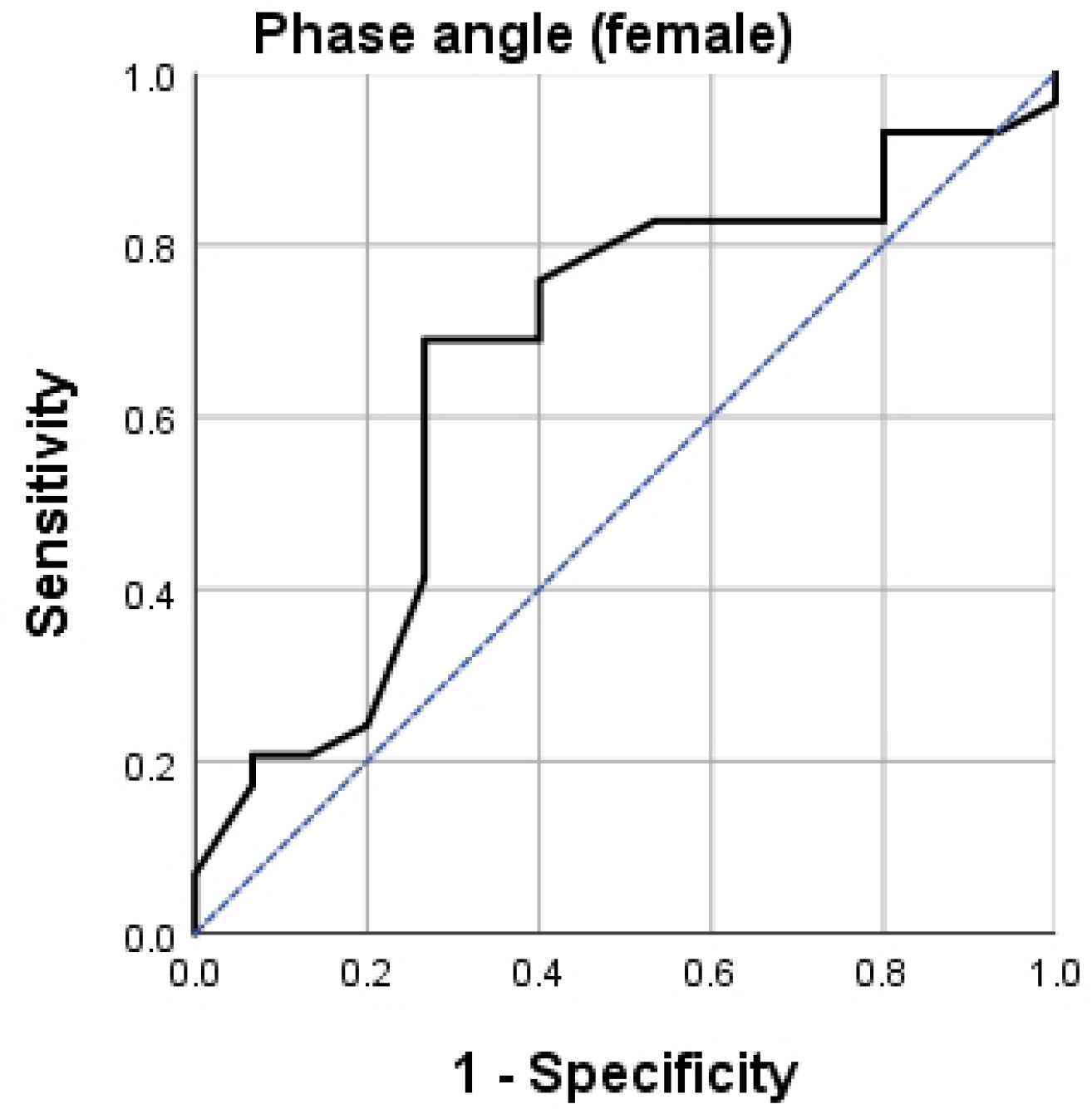
The ROC curve of PhA for sarcopenia in females, the AUCROC is 0.67(p=0.07).

**Fig 3.**
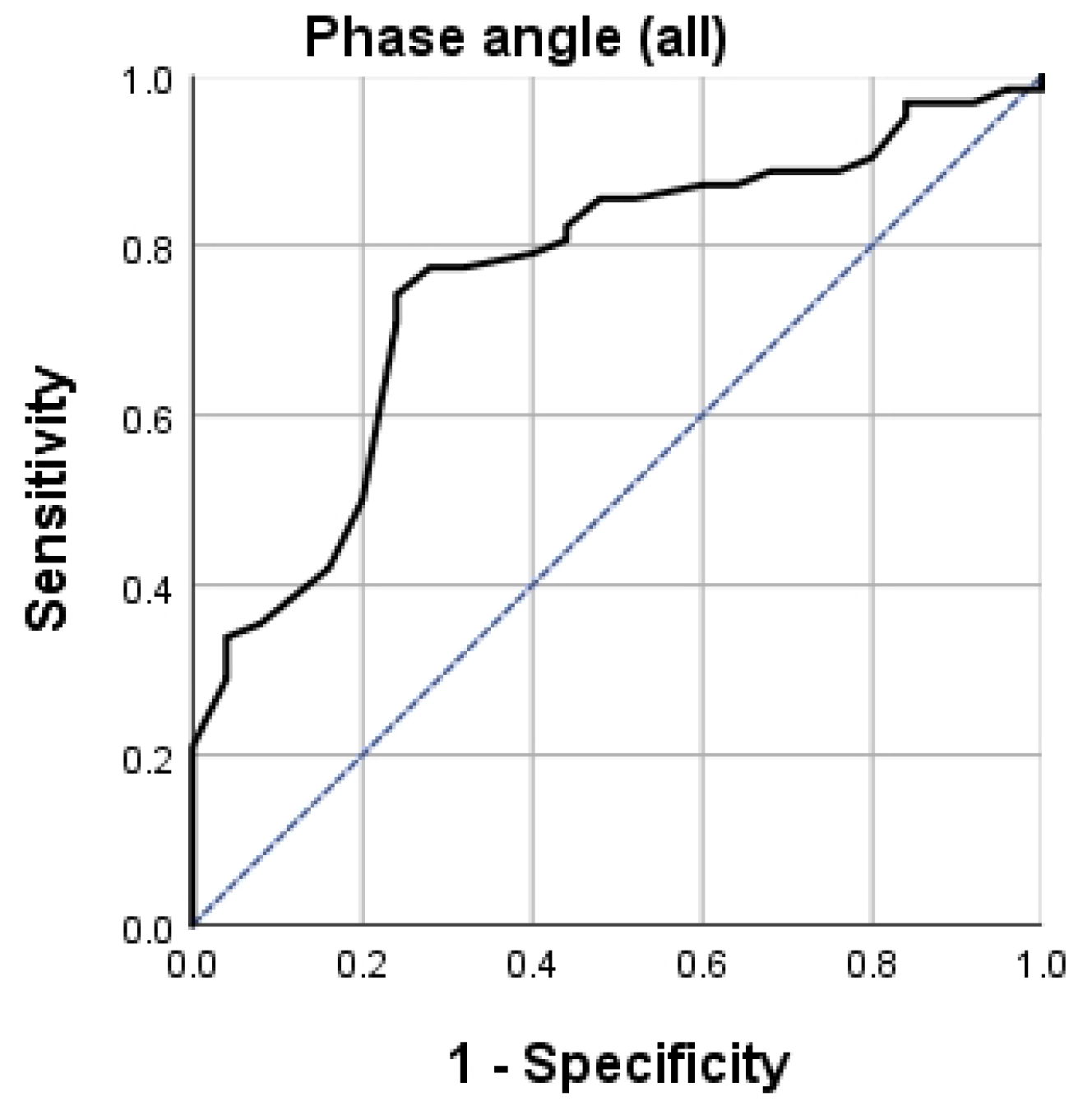
The ROC curve of PhA for sarcopenia in all of the patients, the AUCROC is 0.76(p< 0.01).

## Discussion

Primary sarcopenia is an age-related syndrome characterized by reduced SMM, muscle strength, and muscle function[14], which is associated with various negative outcomes such as falls, disabilities, fractures, and death. Some studies[15, 16] illustrated that the worsened kidney function is associated with an increased prevalence of sarcopenia, especially in dialysis patients. Thus, early diagnosis and intervention of sarcopenia are important for dialysis patients.

The BIA has been used for the last decades for body composition analysis since its simplicity and less harm to the human body[17]. PhA, one of the parameters of BIA, reflects the number of cells and cells’ integrity and has been shown to correlate with nutritional status and sarcopenia. In 2012[18], Marini et al. enrolled 207 elderly and compared the PhA between non-sarcopenia and sarcopenia groups identified by Dual-energy X-ray absorptiometry (DXA). They found PhA was lower in the sarcopenia group and positively correlated with skeletal muscle mass index (SMI). Our study also finds that compared with the non-sarcopenia group, sarcopenia patients have a lower PhA. Meanwhile, in univariate analysis, PhA positively correlated to HGS, SMM, and AMC. But after adjustments for sex, age, diabetes, BMI, Extracellular water ratio, extra water, serum creatinine, total kt/v, and residual kt/v, only HGS and AMC remain associated with PhA. In the multivariate logistic regression analysis, we find no correlation between PhA and sarcopenia. The result is different from the many previous findings. In the elder, <u>Elisabetta Marini</u> et al. found PhA was positively associated with SMI both in male and female(men: r = 0.52, p < 0.01; women: r = 0.31, p < 0.01)[18]. Another study [19] demonstrated a moderate correlation between PhA with HGS. Akihiro et al. revealed that PhA negatively correlated to sarcopenia after a multivariate logistic regression in kidney transplant patients. Han B et al[20] enrolled 80 patients undergoing PD showed that PhA correlated to serum albumin and lean tissue index (LTI) from bioimpedance. Recently, research including 200 patients with PD [21] demonstrated that PhA is independently associated with muscle mass, strength, and sarcopenia, but they used DXA to measure body composition. Whereas, in 2020, Pessoa et al. illustrated that PhA had no association with sarcopenia, SMI, HGS, and walk speed in the old women[22]. Another study in kidney transplant patients[23] also thought PhA was not associated with sarcopenia but with HGS. BIA, consistent with our findings, measured the body composition. Thus, different methods to measure body composition may cause different results between PhA and body composition. Whether PhA is related to sarcopenia and body composition needs more large sample studies.

Upper arm circumference and AMC are both reliable indicators of nutritional status. Still, patients undergoing PD are prone to fat accumulation due to glucose in the peritoneal fluid. Upper arm muscle circumference is a better indicator of nutritional status. An observational cross-sectional study found that mid-arm circumference could act as a predictor of sarcopenia[24]. There are no studies on the relationship between AMC and sarcopenia. To our knowledge, this is the first study to find that PhA correlated to the AMC. However, our method of measuring AMC is BIA rather than the traditional measurement of upper arm circumference and skinfold thickness. The correlation between AMC and PhA needs more research.

We also find that age≥65 is an independent risk factor for sarcopenia, which is consistent with the previous study. Except for age, male gender[16], lower BMI[16, 23], diabetes, longer dialysis duration[24], malnutrition(such as lower serum albumin/pre-albumin)[7, 25], and high-sensitive C-reactive protein(Hs-CRP)[26, 27] were found associated with sarcopenia also in a multivariate logistic regression. Our study also finds serum albumin and pre-albumin are lower in sarcopenia than the non-sarcopenia group, but not sex, BMI, dialysis duration, diabetes, and Hs-CRP. The multivariate regression analysis shows that they are not associated with sarcopenia. This may be caused by the small sample size of our study and the different diagnostic criteria. Meanwhile, after adjustments for other components, the prevalence of sarcopenia is seven times higher in patients with total Kt/V>2.013 than in those with total Kt/V <1.544(OR=7.2, 95% CI:1.635-31.712, P<0.01). A higher Kt/V indicates that the patient is adequately dialyzed, but the dialysis dose is too high. Nutrients such as protein can be lost from the peritoneal fluid, the most important nutrient in muscle synthesis. This may account for the higher risk of sarcopenia in patients with higher Kt/V. This is the first study that demonstrates total Kt/V is a risk factor for sarcopenia, but it is a single-center study, and the result needs to be confirmed by more studies.

In the univariate logistic regression model, PhA correlated to sarcopenia. Thus, we still use ROC curve analysis to find the cut-off value for sarcopenia by phase angle in patients with peritoneal dialysis fluid. We find the best cut-off value of PhA is 5.25°. For males, the best value is PhA≤5.25°and the best value is PhA ≤5.1°for females, which is lower than males. Yamada et al. illustrated that the average PhA for sarcopenia is 4.05°for men and 3.55°for women in community-de-welling Japanese old people[19]. A study[28] on elderly inpatients showed the optimal PhA cut-off value was≤4.55°(sensitivity 70% and specificity 65.9%). In 2019[29], a value≤5.05°was indicated in cirrhosis(sensitivity 71.3% and specificity is 65.5%). Kosoku et al. demonstrated a value of≤4.46 in kidney transplant patients, with a sensitivity of 74% and specificity of 70%[30]. Recently, a study about peritoneal dialysis without peritoneal dialysis fluid showed the cut-off value was≤4.4°(sensitivity 81.3%, specificity 59.6%)[21]. This may be since the total average age of our central dialysis patients(51.3±12.43 year) are lower than that of the patients included in the study (55.5±12.2 year). Except for this, race can also influence PhA. Our study shows a higher cut-off value for males than females, which is consistent with previous studies due to higher muscle mass and lower fat mass in males compared with females.However, whether it can be used as a diagnostic indicator of sarcopenia is related to gender, and its diagnostic significance for male sarcopenia may be greater.

The advantages of our study are the following:1. Several previous studies on sarcopenia had used incomplete criteria, but they used muscle mass alone. Our study uses two variables (HGS and SMI) to diagnose sarcopenia. 2. We use consensus and definitions of sarcopenia for Asia populations.

Our study also has some limitations: 1. This study is a Single-Centre study with a small sample size, and our results still need to be confirmed by large sample, multicenter trials. 2. For the body composition analysis, BIA is used instead of DXA. A guideline suggests that performing would be more reasonable than using a BIA to measure muscle mass in peritoneal dialysis patients.

In conclusion, we find older age and higher total kt/v are risk factors for sarcopenia. PhA is positively associated with HGS and AMC but not with sarcopenia in CAPD patients. These results suggest that PhA can be used as a predictor of muscle mass and strength in CAPD patients, but its’ diagnosis value for sarcopenia needs more studies.

## Data Availability

All relevant data are within the manuscript and its Supporting Information files.

## Acknowledgments

Not applicable.

## Declarations

### Ethics approval and consent to participate

The Ethics Committee approved the study protocol of The Third Hospital of Hebei Medical University (number:20210931). All subjects signed an informed consent form.

### Consent for publication

Not applicable.

### Availability of data and materials

Not applicable.

